# Antiprotozoal medications associated with increased longevity and reduced morbidity in two national cohorts

**DOI:** 10.1101/2025.07.01.25330644

**Authors:** Ariel Israel, Abraham Weizman, Sarah Israel, Shai Ashkenazi, Eytan Ruppin, Shlomo Vinker, Eli Magen, Eugene Merzon

**Author notes:** **Corresponding author:** Ariel Israel, Leumit Research Institute, Sprinzak 23, Tel-Aviv 6473817, Israel.

## Abstract

We conducted a stepwise pharmacoepidemiologic investigation, an unbiased medication-wide screen followed by matched cohort validation and external replication, to identify medications associated with longevity and aging-related morbidity. The initial screen, performed in a large national health system, identified two antiprotozoals, atovaquone-proguanil and mefloquine, as being associated with increased survival. Matched exposed-unexposed cohorts were then constructed to validate mortality associations and examine incident outcomes, revealing reduced risks for diabetes, dementia, cardiovascular, renal, hepatic, pulmonary, and selected cancers, alongside increased risks for hearing loss, dry eye/Sjogren’s, and lichen planus. These patterns were externally replicated in the US TriNetX network, where similar associations were observed for atovaquone-proguanil, mefloquine, and nirmatrelvir-ritonavir. Because nirmatrelvir-ritonavir is prescribed to older, multimorbid individuals, its associations are unlikely to reflect healthy-traveler conditioning. The concordant protective and tissue-specific adverse associations across datasets and antiprotozoal drug classes support a testable hypothesis: short antiprotozoal courses may reduce aging-related morbidity by decreasing persistent protozoal burden, particularly *Toxoplasma gondii*.

## Introduction

Identifying the long-term effects of medications is a central goal of pharmacoepidemiology and drug repurposing research. Many approved drugs may exert previously unrecognized influences on human health, with potential benefits or harms extending beyond their original indications. Medications that promote healthy aging would be expected to reduce mortality and lower the incidence of chronic disease.

Electronic health records (EHRs) from large healthcare organizations offer a powerful resource for systematically evaluating the long-term effects of medications on mortality and age-related morbidity ^1–3^. Using EHR data from Leumit Health Services (LHS), a nationwide provider in Israel, we performed a medication-wide exploratory screen to identify drugs associated with increased longevity. This discovery analysis highlighted two antiprotozoal agents, atovaquone-proguanil and mefloquine, both primarily prescribed for malaria prophylaxis in travelers. These medications have not previously been implicated in long-term survival or aging-related morbidity. Their emergence from an unbiased, medication-wide screen motivated the targeted evaluation of these medications, which were selected to address an apparent knowledge gap rather than through hypothesis-driven preselection.

The identification of antiprotozoal medications as longevity-associated agents is consistent with emerging evidence linking the human microbiome to age-related diseases, including cardiovascular ^4^, metabolic ^5^, malignant ^6^ and neurodegenerative disorders ^7,8^. Antiprotozoal agents may modulate the microbiome—particularly through elimination of specific protozoal pathogens—and thereby influence long-term health outcomes beyond their original therapeutic purposes.

To explore and validate these findings, we undertook a sequence of follow-up analyses. In LHS, we constructed matched exposed-unexposed cohorts to validate mortality associations and to examine the incidence of common age-related conditions in the decade following the first uptake of antiprotozoal medications. We then sought external validation in the US-based TriNetX federated network, comprising EHR data from more than 120 million individuals. There, we examined atovaquone-proguanil and mefloquine, prescribed mainly for malaria prophylaxis in travelers, alongside nirmatrelvir-ritonavir, an antiviral used in older and comorbid patients at risk for COVID-19 complications that also has antiprotozoal activity. Inclusion of this latter cohort helped address potential “healthy traveler” bias by confirming that protective associations extend to non-traveler populations. Across these analyses, we observed consistent associations between antiprotozoal exposure and reduced mortality, accompanied by concordant changes in the incidence of multiple age-related diseases.

**Figure 1** summarizes the study design, which comprised three sequential analyses: a discovery screen in LHS (Analysis A), a validation-exploration cohort in LHS (Analysis B), and external validation in TriNetX (Analysis C).

**Figure 1.**
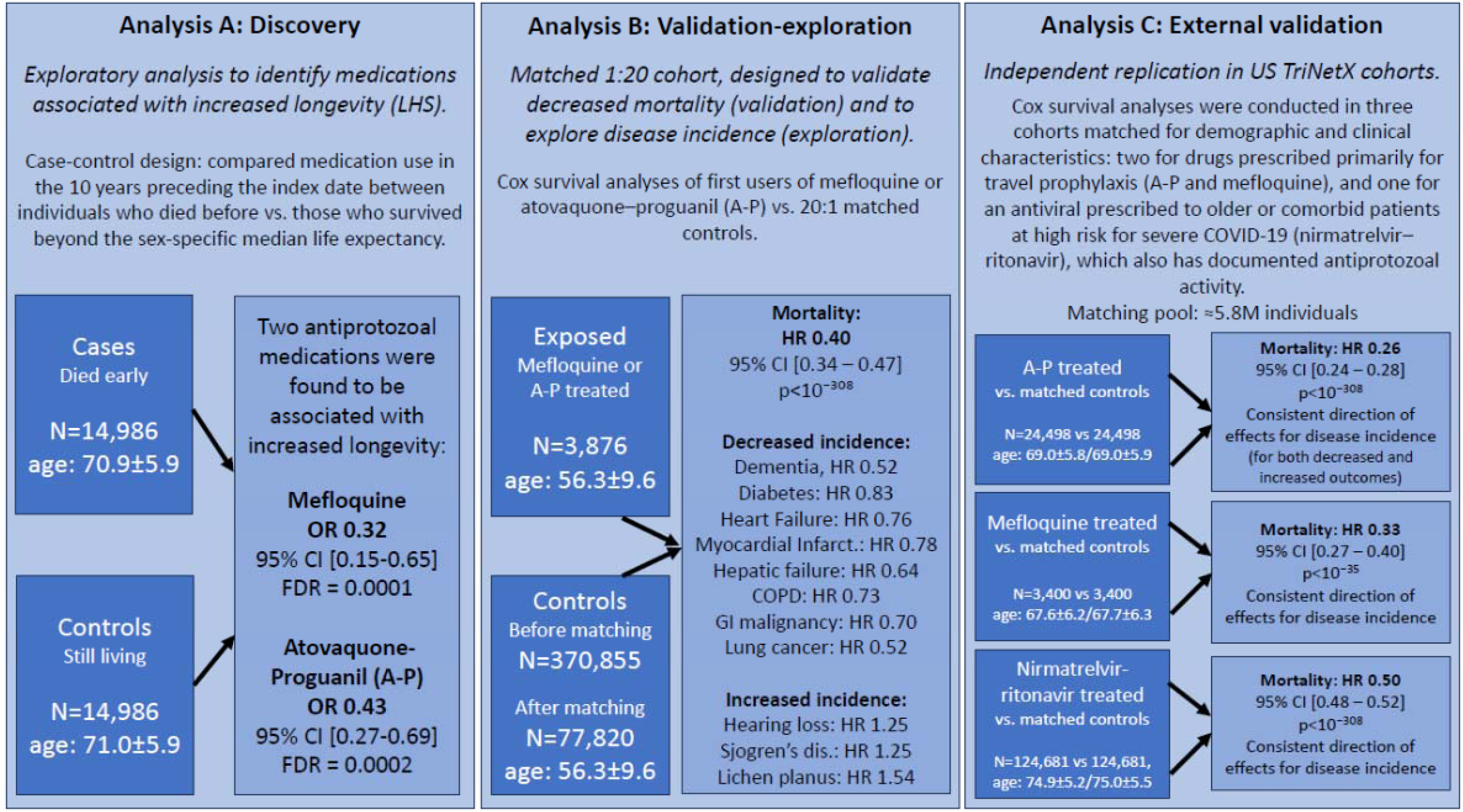
Design of the analyses performed to identify and validate medications associated with increased longevity and decreased age-related morbidity.

## Results

### Analysis A: LHS exploratory cohort

Table 1 summarizes the demographic and clinical characteristics of 14,986 patients who died before the cohort median age of death (cases) and 14,986 matched survivors (controls) in the LHS cohort. Matching ensured similar distributions of age, sex, and socioeconomic status (49.2% female, mean age 71.0 ± 5.9 years at index). Compared to controls, cases had slightly higher BMI (29.1 ± 6.0 vs. 28.5 ± 4.8 kg/m²) and weight (77.5 ± 17.7 vs. 76.3 ± 14.6 kg) and were more likely to be smokers (OR 2.00, 95% CI 1.86-2.15). After an average follow-up of 12.5 years, controls reached a mean age of 83.6 ± 5.9 years, while cases died at a mean age of 74.3 ± 5.7 years (Table 1B).

**Table 1.**
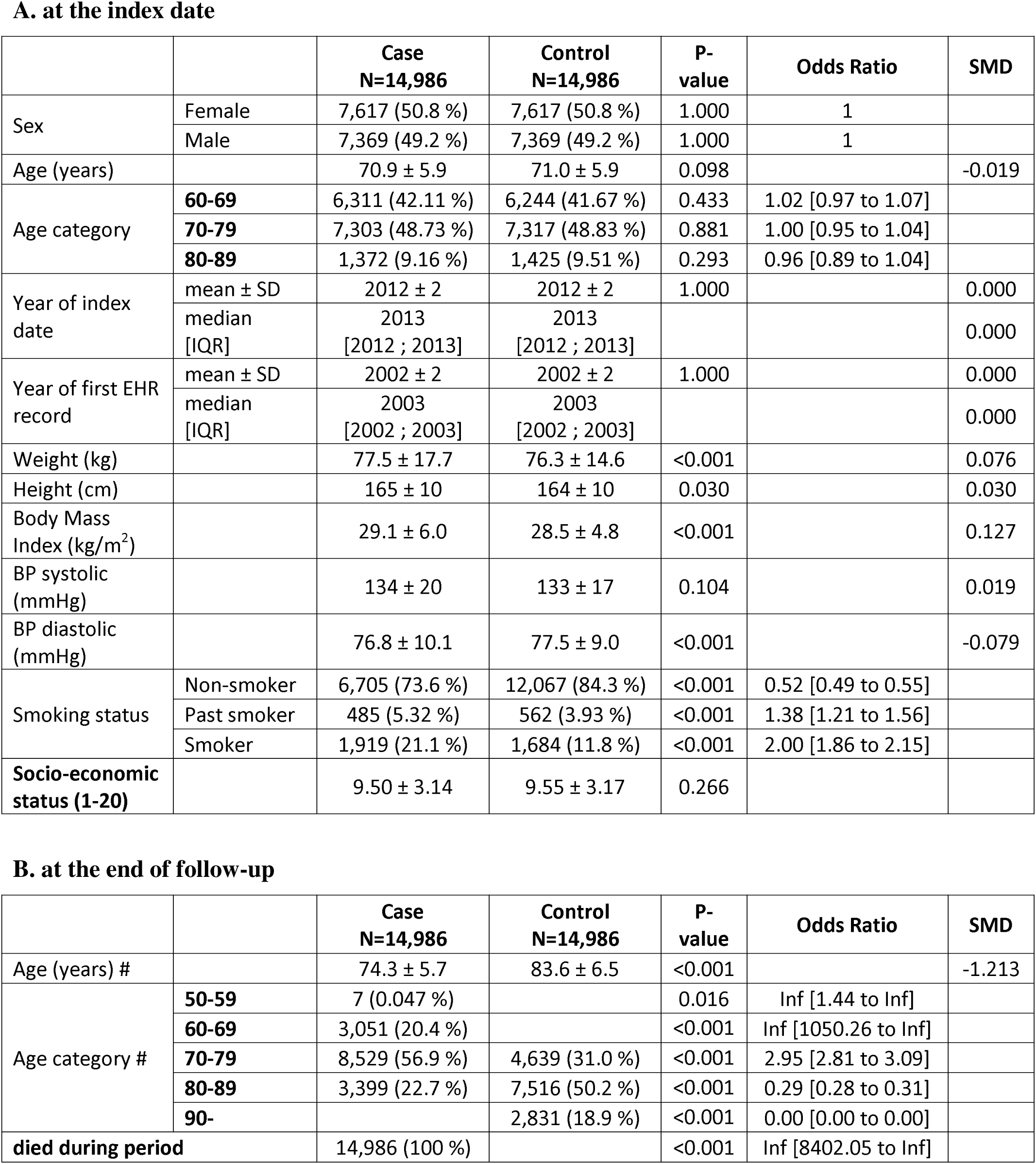
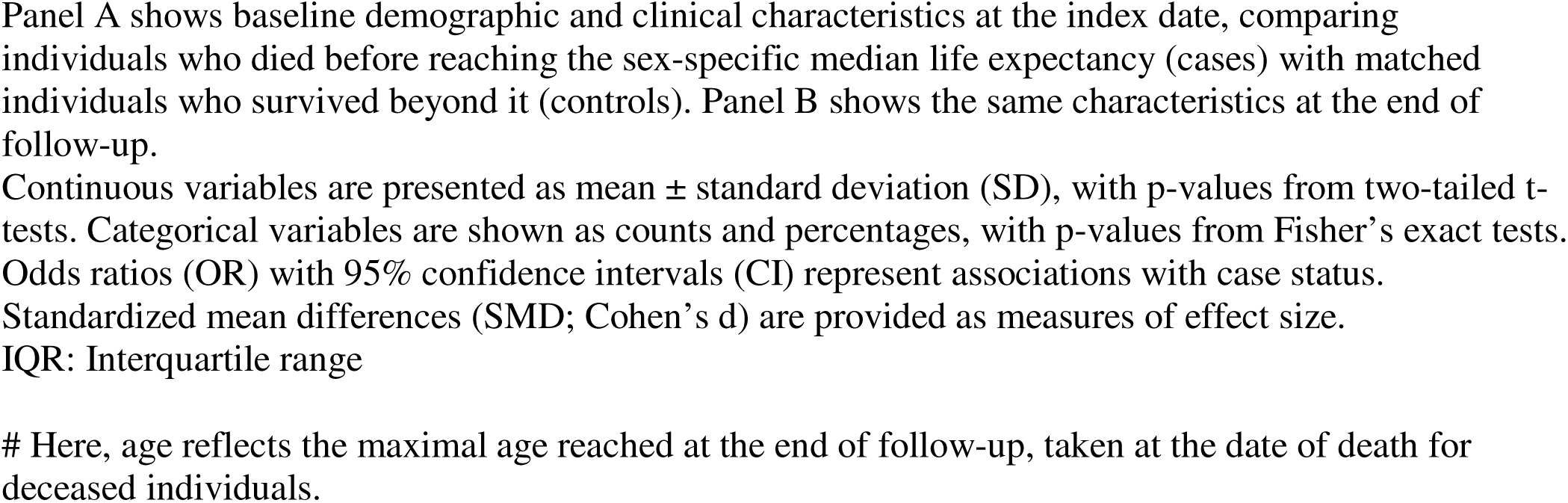
Demographic and clinical characteristics of the LHS exploratory longevity cohort (Analysis A).

### Analysis A results: Medications associated with decreased mortality

Medication use during the preceding decade was systematically evaluated for associations with mortality. Out of 1470 screened compounds, seven agents, from diverse therapeutic classes met the prespecified OR and FDR threshold (OR<0.5, FDR <0.5%, Benjamini-Hochberg correction). The ATC class P01B (antimalarials) was the only therapeutic category represented by multiple top entries, prompting downstream validation of these two antiprotozoal agents: mefloquine (OR 0.32, 95% CI 0.15–0.65, FDR = 0.0001) and atovaquone–proguanil (OR 0.43, 95% CI 0.27–0.69, FDR = 0.0002) (Supplementary Table 1).

### Analysis B: Validation-Exploration Cohort in LHS

These findings prompted a more detailed evaluation of mortality and morbidity outcomes in matched cohorts. We constructed a cohort of individuals aged 40-85 who had received at least one prescription for either A-P or mefloquine, and matched each treated patient to 20 unexposed controls based on sex, socioeconomic status (SES), and year of first EHR record. The final cohort included 3,876 treated individuals and 77,820 controls with broadly comparable baseline characteristics (Table 2).

**Table 2.** Demographic and clinical characteristics of the LHS cohort of individuals treated with atovaquone-proguanil or mefloquine versus matched controls (Analysis B).

|  |  | Case<br>N=3,876 | Control<br>N=77,820 | P-value | Odds Ratio | SMD |
| --- | --- | --- | --- | --- | --- | --- |
| Sex | Female | 2,028 (52.3 %) | 40,560 (52.3 %) | 1.000 | 1 |  |
|  | Male | 1,848 (47.7 %) | 36,960 (47.7 %) | 1.000 | 1 |  |
| Age at index (years) |  | 56.3 ± 9.4 | 56.3 ± 9.6 | 0.990 | 1.02 [0.95 to 1.10] | 0.000 |
| Age category | **40-49** | 1,100 (28.4 %) | 21,624 (27.9 %) | 0.509 | 0.92 [0.86 to 0.99] |  |
|  | **50-59** | 1,325 (34.2 %) | 27,903 (36.0 %) | 0.023 | 1.11 [1.03 to 1.19] |  |
|  | **60-69** | 1,111 (28.7 %) | 20,594 (26.6 %) | 0.004 | 1.02 [0.91 to 1.15] |  |
|  | **70-79** | 326 (8.41 %) | 6,389 (8.24 %) | 0.697 | 0.27 [0.15 to 0.46] |  |
|  | **80-89** | 14 (0.36 %) | 1,010 (1.30 %) | <0.001 | 1.02 [0.95 to 1.10] |  |
| Year of index date | mean ± SD | 2015 ± 4 | 2015 ± 4 | 1.000 |  | 0.000 |
|  | median [IQR] | 2016 [2013 ; 2018] | 2016 [2013 ; 2018] |  |  | 0.000 |
| Weight (kg) |  | 76.3 ± 15.3 | 77.3 ± 16.5 | <0.001 |  | -0.060 |
| Height (cm) |  | 168 ± 9 | 167 ± 10 | <0.001 |  | 0.079 |
| Body Mass Index (kg/m<sup>2</sup>) |  | 27.0 ± 4.5 | 27.5 ± 5.2 | <0.001 |  | -0.114 |
| BP systolic (mmHg) |  | 134 ± 20 | 133 ± 17 | 0.104 |  | 0.019 |
| BP diastolic (mmHg) |  | 76.8 ± 10.1 | 77.5 ± 9.0 | <0.001 |  | -0.079 |
| Smoking status | Non-smoker | 3,168 (82.8 %) | 56,248 (75.3 %) | <0.001 | 1.58 [1.45 to 1.73] |  |
|  | Past smoker | 105 (2.74 %) | 2,181 (2.92 %) | 0.587 | 0.94 [0.76 to 1.15] |  |
|  | Smoker | 553 (14.5 %) | 16,301 (21.8 %) | <0.001 | 0.61 [0.55 to 0.66] |  |
| **Ethnic sector** | Arab | 88 (2.27 %) | 1,760 (2.27 %) | 1.000 | 1 |  |
|  | General | 3,591 (92.7 %) | 71,820 (92.7 %) | 1.000 | 1 |  |
|  | Jewish ultra-orthodox | 197 (5.1 %) | 3,940 (5.1 %) | 1.000 | 1 |  |

Baseline demographic and clinical characteristics of Leumit Health Services (LHS) patients exposed to atovaquone-proguanil or mefloquine (cases) compared with matched controls. Matching was performed on birth year, sex, ethnic sector, and socioeconomic status (SES) category. Reported values reflect characteristics at the index date (first prescription for cases; assigned index date for controls). Counts and percentages represent the distribution of categorical variables in cases and controls at baseline. Odds ratios (OR) with 95% confidence intervals (CI) are shown for categorical comparisons, alongside p-values from Fisher’s exact test. Continuous variables are presented as mean ± standard deviation (SD), with standardized mean differences (SMD; Cohen’s d) provided as effect size estimates. Abbreviations: BP, blood pressure; BMI, body mass index.

The mean age at index date was 56.3 years in both groups, with 52.3% female participants. Small but statistically significant baseline differences were noted for BMI (27.0 ± 4.5 vs. 27.5 ± 5.2 kg/m², p < 0.001), weight (76.3 ± 15.3 vs. 77.3 ± 16.5 kg, p < 0.001), and height (168 ± 9 vs. 167 ± 10 cm, p < 0.001). Systolic blood pressure was similar across groups, while diastolic blood pressure was slightly lower among treated individuals (76.8 ± 10.1 vs. 77.5 ± 9.0 mmHg, p < 0.001). Current smoking was significantly less frequent in the treated group (14.5% vs. 21.8%, p < 0.001), although past smoking rates and ethnic sector distributions were similar between cases and controls.

### Analysis B: Cox proportional hazards models

We next evaluated the association between exposure to A-P or mefloquine and subsequent mortality and morbidity using Cox proportional hazards models (cumulative incidence plots in Figure 2). Treated individuals had markedly reduced risk of all-cause mortality (HR = 0.40 [95% CI, 0.34-0.47], p < 0.001; Figure 2A). Significant reductions were also observed for dementia (HR = 0.52 [0.41-0.67], p < 0.001; Figure 2B), diabetes mellitus (HR = 0.83 [0.75-0.92], p = 0.0006; Figure 2C), heart failure (HR = 0.76 [0.63-0.91], p = 0.006; Figure 2D), myocardial infarction (HR = 0.78 [0.66-0.92], p = 0.003; Figure 2E), hepatic failure (HR = 0.64 [0.43-0.95], p = 0.03; Figure 2F), chronic obstructive pulmonary disease (COPD) (HR = 0.73 [0.62-0.86], p = 0.0002; Figure 2G), gastrointestinal malignancy (HR = 0.70 [0.52-0.96], p = 0.03; Figure 2H), and lung cancer (HR = 0.52 [0.33-0.82], p = 0.004; Figure 2I).

**Figure 2.**
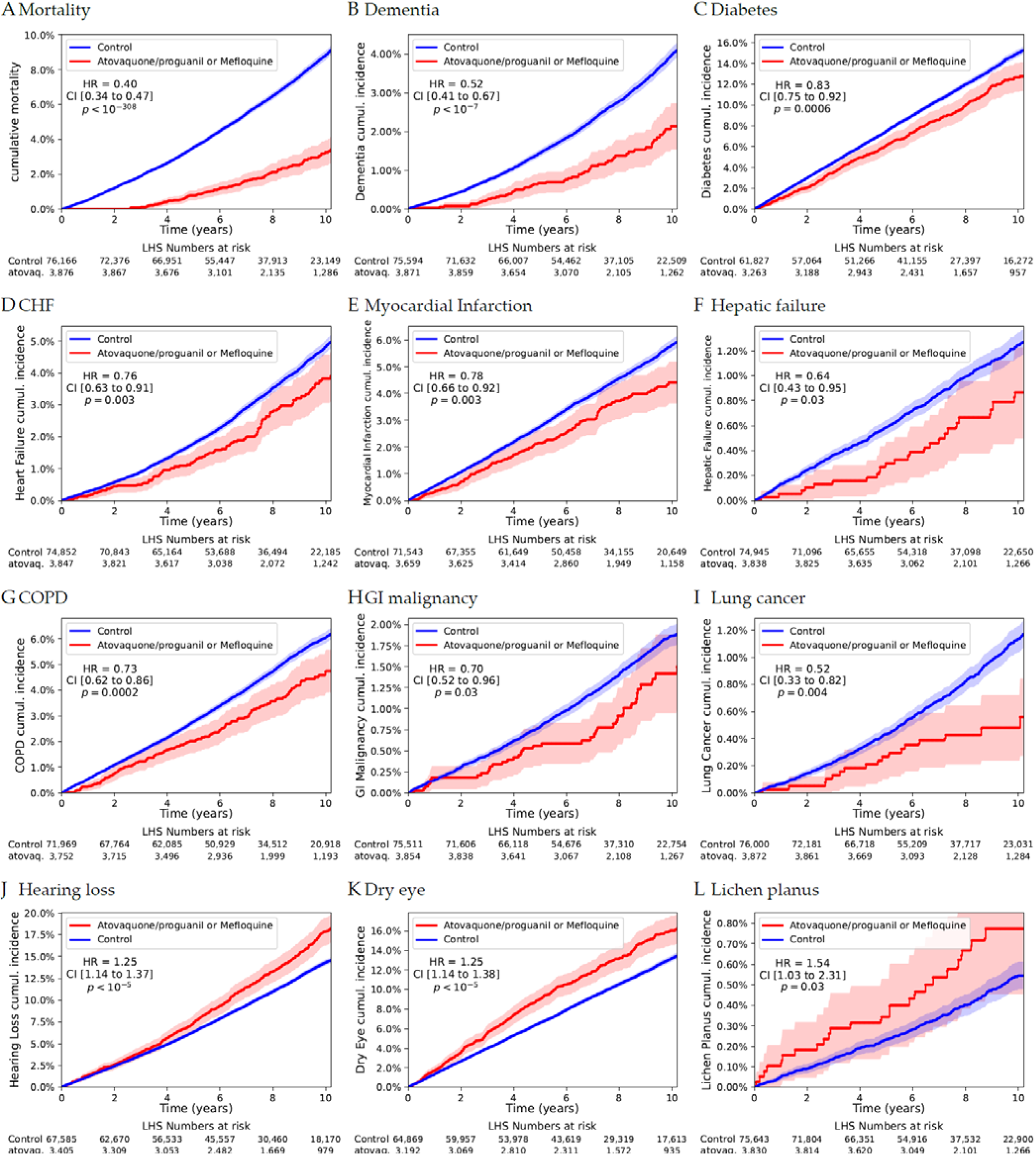
Cumulative mortality and incidence of major health outcomes in patients treated with antiprotozoals versus matched controls in LHS (Analysis B). Kaplan-Meier curves showing cumulative incidence of major health outcomes among Leumit Health Services (LHS) patients exposed to mefloquine or atovaquone-proguanil (red) compared with matched controls (blue). Hazard ratios (HR), 95% confidence intervals (CI), and p-values were estimated from Cox proportional hazards models. Follow-up was censored at the date of death or last recorded visit, whichever occurred later, and individuals with a diagnosis of the outcome prior to the index date were excluded. Shaded areas represent 95% CI. Panel labels: A, all-cause mortality; B, dementia; C, diabetes mellitus; D, congestive heart failure (CHF); E, myocardial infarction; F, hepatic failure; G, chronic obstructive pulmonary disease (COPD); H, gastrointestinal (GI) malignancy; I, lung malignancy; J, hearing loss; K, dry eye; L, lichen planus.

Conversely, treated individuals exhibited significantly increased risks of hearing loss (HR = 1.25 [1.14-1.37], p < 0.001; Figure 2J), Sjögren’s disease/eye dryness (HR = 1.25 [1.14-1.38], p < 0.001; Figure 2K), and lichen planus (HR = 1.54 [1.03-2.31], p = 0.03; Figure 2L).

A forest plot summarizing these outcomes (Figure 3) illustrates the broad reductions in mortality and major age-related conditions associated with antiprotozoal exposure, alongside increased risks of specific auditory, ophthalmic, and dermatologic conditions.

**Figure 3.**
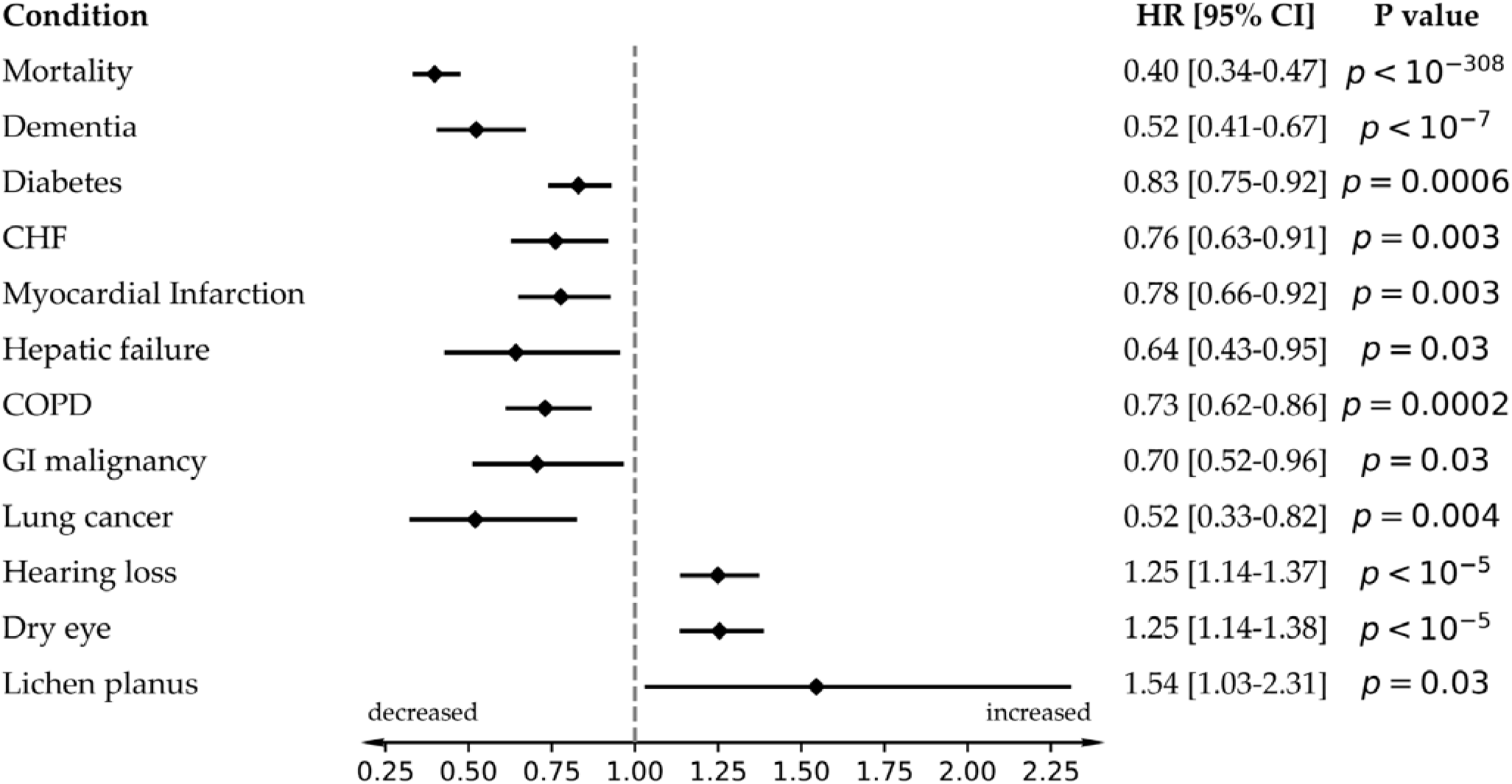
Forest plot of selected health outcomes in patients treated with atovaquone-proguanil or mefloquine vs. matched controls in LHS (Analysis B). Hazard ratios (HR) and 95% confidence intervals (CI) for all-cause mortality and selected age-related conditions among patients treated with mefloquine or atovaquone-proguanil, compared with matched controls in Leumit Health Services (LHS). Outcomes were estimated using Cox proportional hazards models, with follow-up censored at the date of death or last recorded visit, whichever occurred later. Individuals with a diagnosis of the respective condition prior to the index date were excluded. Horizontal bars indicate 95% CIs; values to the left of 1 indicate reduced risk, and those to the right indicate increased risk. Protective associations were identified for mortality, dementia, diabetes, cardiovascular events, hepatic failure, COPD, and several malignancies. Conversely, increased risks were observed for hearing loss, dry eye, and lichen planus.

Outcomes that demonstrated statistically significant increases or decreases in LHS were retained for evaluation in TriNetX, irrespective of effect direction. These patterns of both reduced and increased risk therefore motivated the specific set of outcomes selected for replication.

### Analysis C: Validation cohorts in the US-based TrinetX collaborative network

To validate the associations observed in the LHS national cohort, we conducted independent analyses in TriNetX, a federated network comprising EHRs from over 70 US healthcare organizations and more than 120 million patients. Eligible individuals were those born 70–89 years before the cohort extraction date, corresponding to individuals who would be aged 70–89 at extraction, irrespective of vital status. Cases were defined as those with a record of the medication of interest, while controls had no such exposure. Propensity score matching (PSM) was applied to balance groups on age, sex, race, ethnicity, smoking status, diabetes, BMI, and HbA1c at baseline.

To address potential indication bias stemming from the use of antimalarials primarily in travelers, we included a third validation cohort exposed to nirmatrelvir-ritonavir (N-R, Paxlovid). This drug was selected because of its predicted antiprotozoal properties and its widespread prescription as an antiviral during the COVID-19 pandemic. The N-R cohort offered substantially greater statistical power than the smaller A-P and mefloquine cohorts; however, follow-up was limited to a maximum of three years given the recent introduction of this medication, in contrast to the longer observation periods available for A-P and mefloquine.

The final matched cohorts included 24,498 exposed individuals and 24,498 controls for A-P, 3,400 exposed and 3,400 controls for mefloquine, and 124,681 exposed and 124,681 controls for N-R (Table 3). Across cohorts, mean age at index was closely matched (69.0 ± 5.8 years for A-P, 67.6 ± 6.2 years for mefloquine, and 75.0 ± 5.5 years for N-R). The close equivalence of both age at index and current age after matching empirically confirms identical follow-up time windows in exposed and unexposed groups, supporting the absence of immortal-time and calendar bias. Sex distributions were balanced (55% female for A-P, 52% for mefloquine, 58% for N-R). Race and ethnicity distributions were similar within each matched cohort, with the majority identified as White (83% for A-P, 56% for mefloquine, 85% for N-R). Baseline prevalence of nicotine dependence (3-8%), diabetes mellitus (14-26%), and overweight/obesity (13-27%) were comparable between groups. HbA1c and BMI were also closely matched, ensuring well-balanced baseline metabolic profiles for subsequent outcome analyses.

**Table 3.**
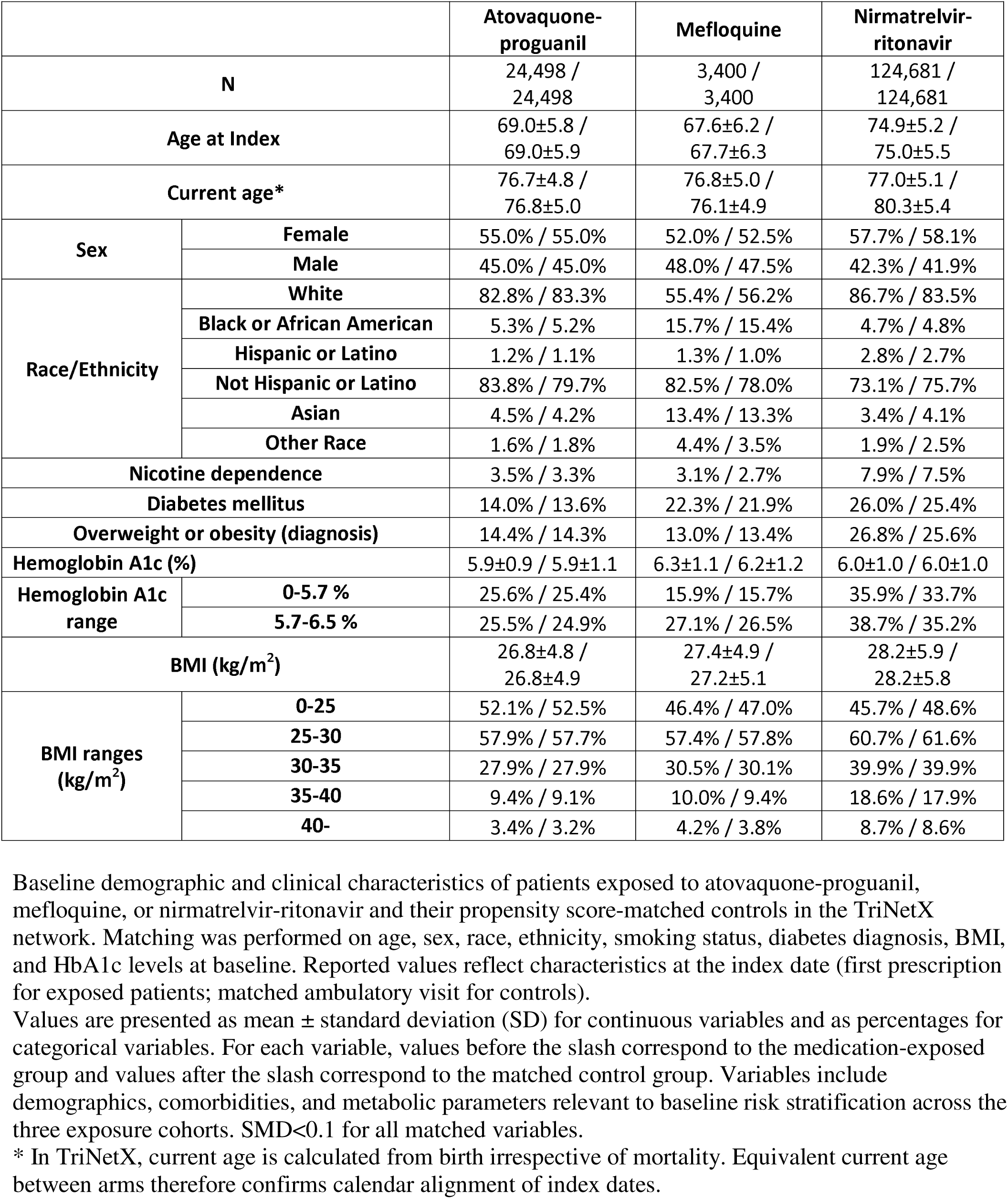
Demographic and clinical characteristics of the matched TriNetX cohorts for atovaquone-proguanil, mefloquine, and nirmatrelvir-ritonavir exposure (Analysis C).

|  |  | Atovaquone-proguanil | Mefloquine | Nirmatrelvir-ritonavir |
| --- | --- | --- | --- | --- |
| **N** |  | 24,498 / 24,498 | 3,400 / 3,400 | 124,681 / 124,681 |
| **Age at Index** |  | 69.0±5.8 / 69.0±5.9 | 67.6±6.2 / 67.7±6.3 | 74.9±5.2 / 75.0±5.5 |
| **Current age\*** |  | 76.7±4.8 / 76.8±5.0 | 76.8±5.0 / 76.1±4.9 | 77.0±5.1 / 80.3±5.4 |
| **Sex** | **Female** | 55.0% / 55.0% | 52.0% / 52.5% | 57.7% / 58.1% |
|  | **Male** | 45.0% / 45.0% | 48.0% / 47.5% | 42.3% / 41.9% |
| **Race/Ethnicity** | **White** | 82.8% / 83.3% | 55.4% / 56.2% | 86.7% / 83.5% |
|  | **Black or African American** | 5.3% / 5.2% | 15.7% / 15.4% | 4.7% / 4.8% |
|  | **Hispanic or Latino** | 1.2% / 1.1% | 1.3% / 1.0% | 2.8% / 2.7% |
|  | **Not Hispanic or Latino** | 83.8% / 79.7% | 82.5% / 78.0% | 73.1% / 75.7% |
|  | **Asian** | 4.5% / 4.2% | 13.4% / 13.3% | 3.4% / 4.1% |
|  | **Other Race** | 1.6% / 1.8% | 4.4% / 3.5% | 1.9% / 2.5% |
| **Nicotine dependence** |  | 3.5% / 3.3% | 3.1% / 2.7% | 7.9% / 7.5% |
| **Diabetes mellitus** |  | 14.0% / 13.6% | 22.3% / 21.9% | 26.0% / 25.4% |
| **Overweight or obesity (diagnosis)** |  | 14.4% / 14.3% | 13.0% / 13.4% | 26.8% / 25.6% |
| **Hemoglobin A1c (%)** |  | 5.9±0.9 / 5.9±1.1 | 6.3±1.1 / 6.2±1.2 | 6.0±1.0 / 6.0±1.0 |
| **Hemoglobin A1c range** | **0-5.7 %** | 25.6% / 25.4% | 15.9% / 15.7% | 35.9% / 33.7% |
|  | **5.7-6.5 %** | 25.5% / 24.9% | 27.1% / 26.5% | 38.7% / 35.2% |
| **BMI (kg/m<sup>2</sup>)** |  | 26.8±4.8 / 26.8±4.9 | 27.4±4.9 / 27.2±5.1 | 28.2±5.9 / 28.2±5.8 |
| **BMI ranges (kg/m<sup>2</sup>)** | **0-25** | 52.1% / 52.5% | 46.4% / 47.0% | 45.7% / 48.6% |
|  | **25-30** | 57.9% / 57.7% | 57.4% / 57.8% | 60.7% / 61.6% |
|  | **30-35** | 27.9% / 27.9% | 30.5% / 30.1% | 39.9% / 39.9% |
|  | **35-40** | 9.4% / 9.1% | 10.0% / 9.4% | 18.6% / 17.9% |
|  | **40-** | 3.4% / 3.2% | 4.2% / 3.8% | 8.7% / 8.6% |

Baseline demographic and clinical characteristics of patients exposed to atovaquone-proguanil, mefloquine, or nirmatrelvir-ritonavir and their propensity score-matched controls in the TriNetX network. Matching was performed on age, sex, race, ethnicity, smoking status, diabetes diagnosis, BMI, and HbA1c levels at baseline. Reported values reflect characteristics at the index date (first prescription for exposed patients; matched ambulatory visit for controls). Values are presented as mean ± standard deviation (SD) for continuous variables and as percentages for categorical variables. For each variable, values before the slash correspond to the medication-exposed group and values after the slash correspond to the matched control group. Variables include demographics, comorbidities, and metabolic parameters relevant to baseline risk stratification across the three exposure cohorts. SMD<0.1 for all matched variables. \* In TriNetX, current age is calculated from birth irrespective of mortality. Equivalent current age between arms therefore confirms calendar alignment of index dates.

### Analysis C: Cox proportional hazard models

In the TriNetX cohorts, we evaluated mortality and age-related conditions identified in the LHS analyses, excluding individuals with a prior diagnosis of each outcome at baseline. Cumulative incidence plots are presented in Figure 4.

**Figure 4.**
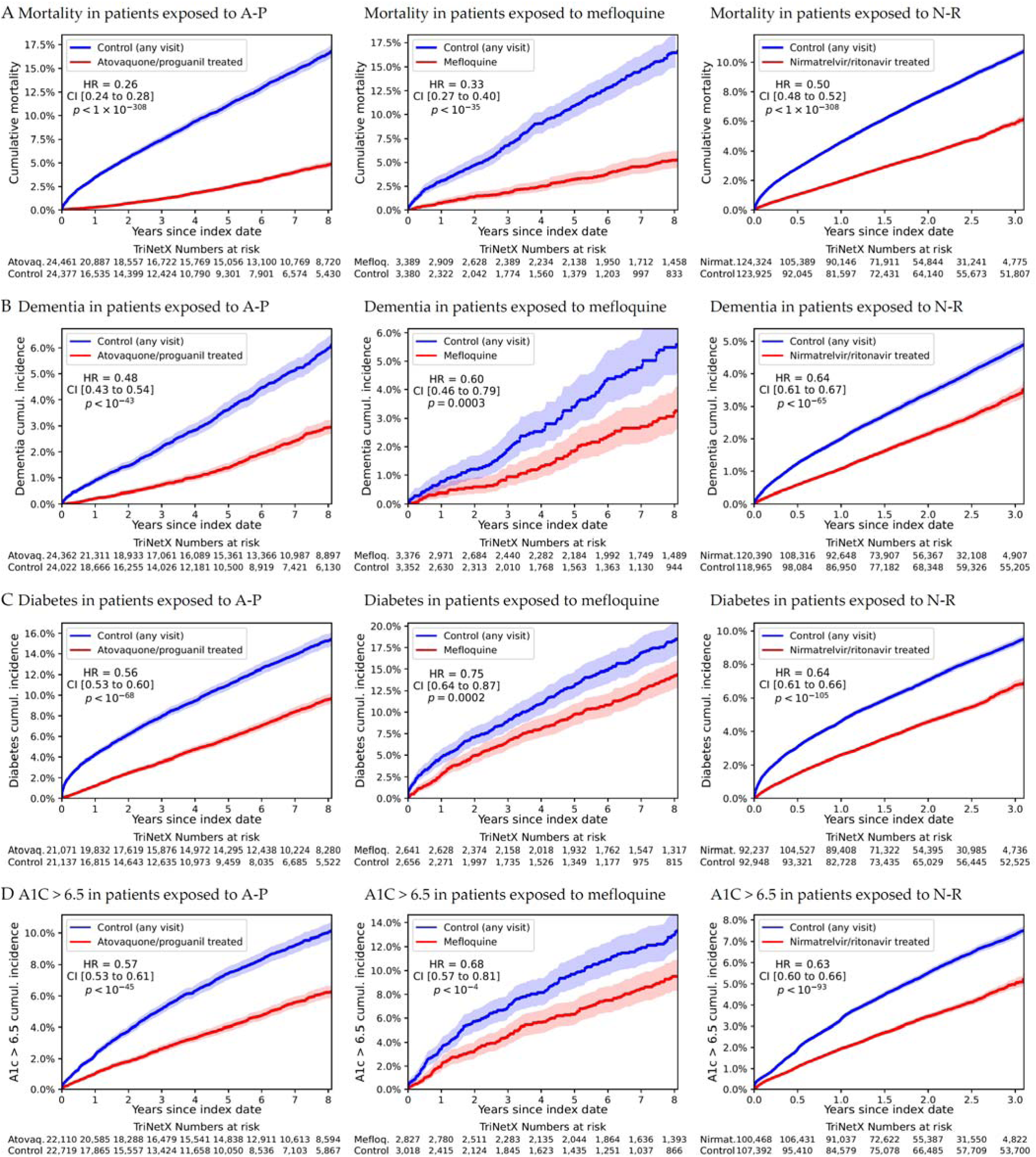
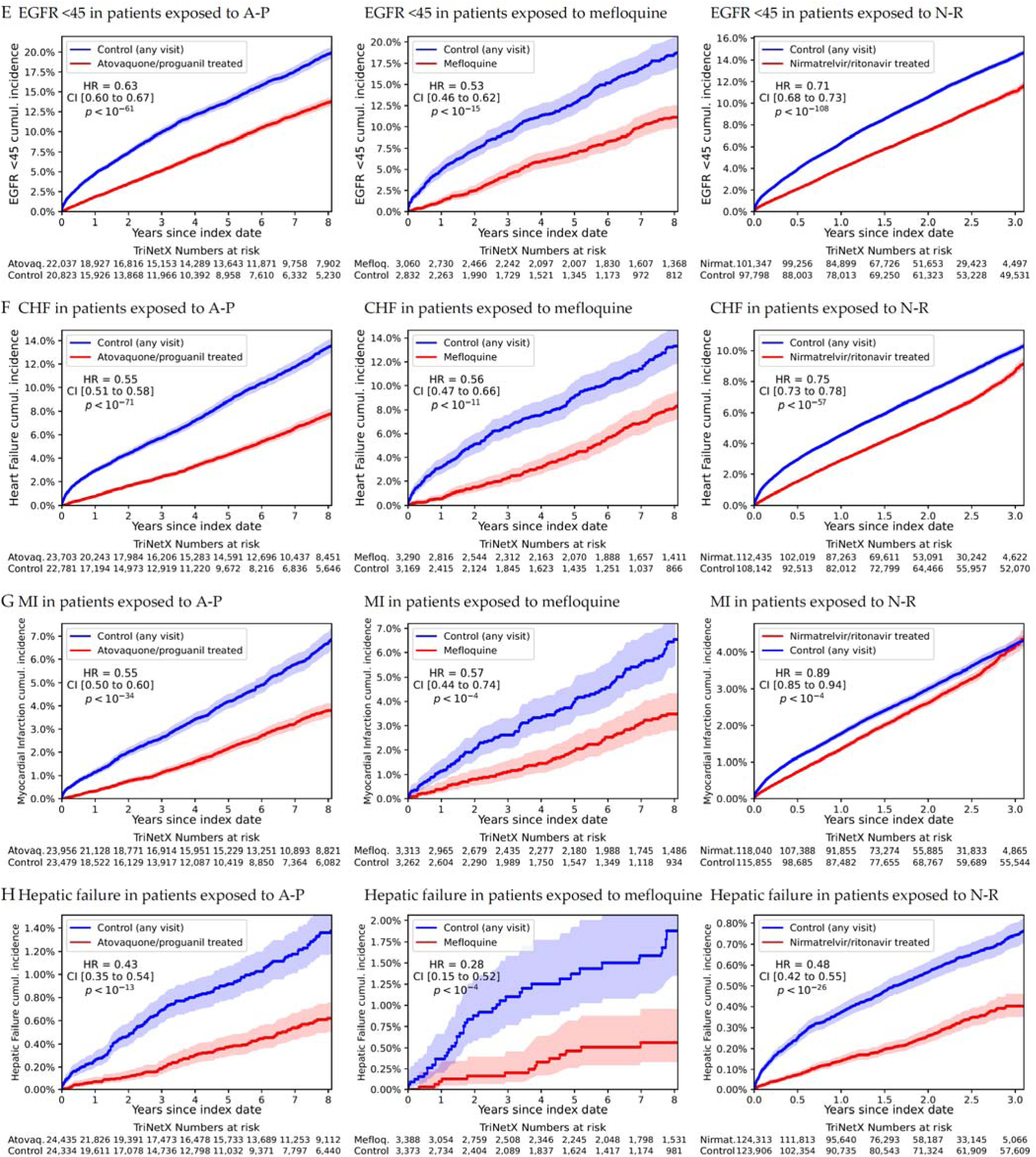
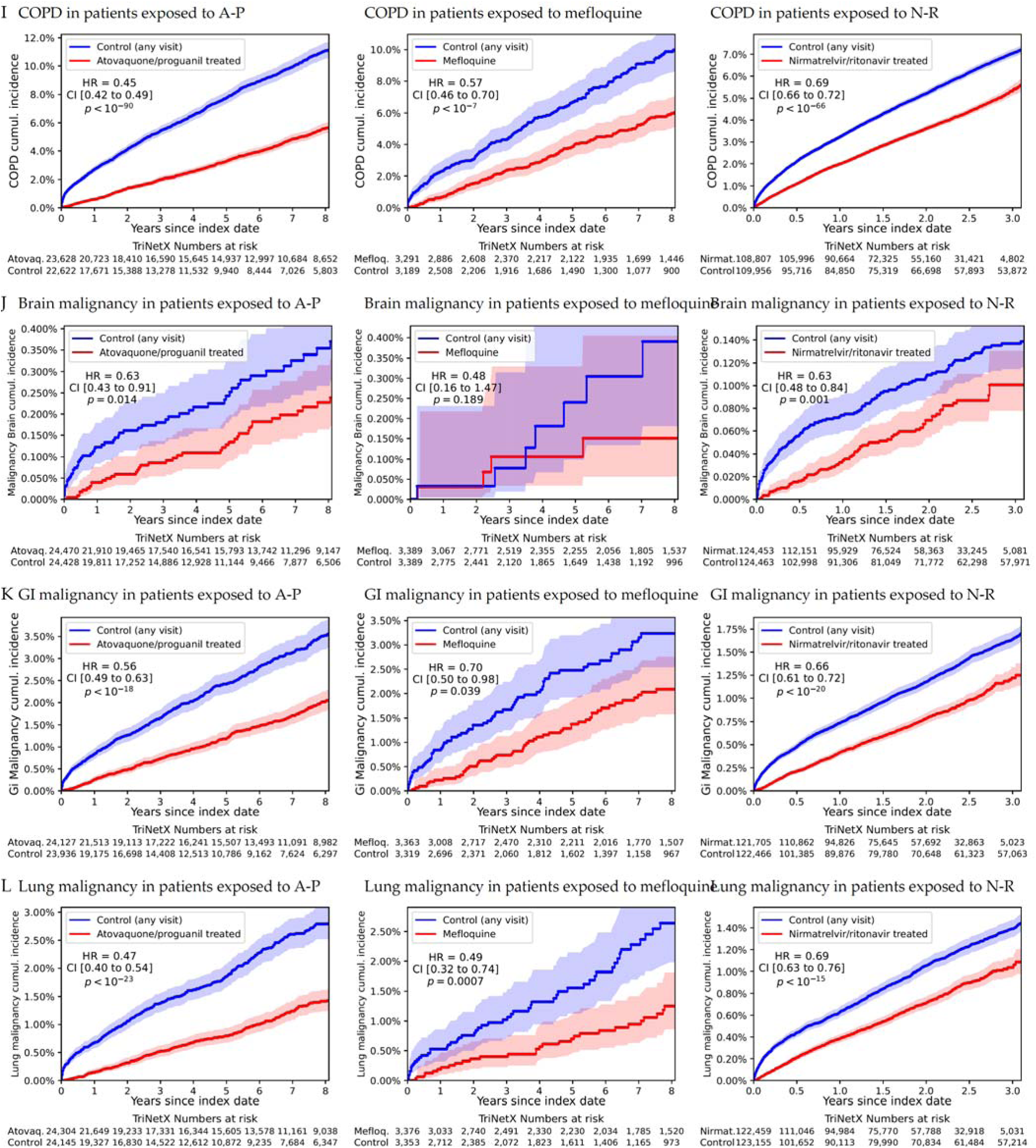
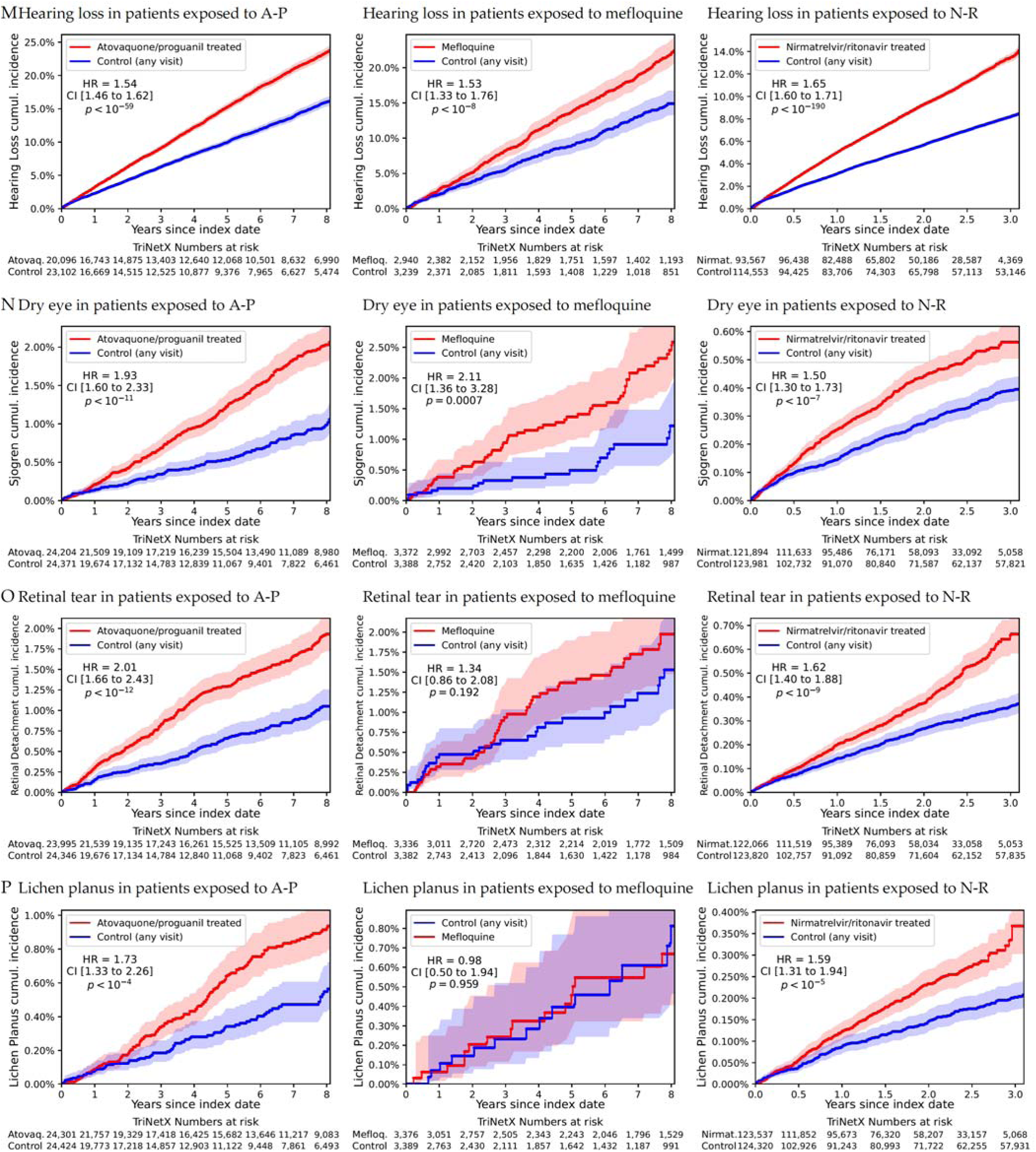
Cumulative incidence of mortality and age-related health outcomes following antiprotozoal exposure in TriNetX cohorts (Analysis C). Cumulative incidence curves comparing patients treated with atovaquone-proguanil (left), mefloquine (middle), and nirmatrelvir-ritonavir (right) versus propensity score-matched controls in the TriNetX network. Outcomes include all-cause mortality and selected age-related conditions identified in the LHS cohort, with follow-up extending up to eight years for atovaquone-proguanil and mefloquine and up to three years for nirmatrelvir-ritonavir. Individuals with prior diagnoses of the respective outcomes at baseline were excluded from each analysis. Hazard ratios (HR), 95% confidence intervals (CI), and p-values were estimated using Cox proportional hazards models. Associations included consistent reductions in mortality and morbidity across multiple outcomes, while increased risks were identified for certain adverse events, including hearing loss and ophthalmic conditions.

Consistent and significant reductions in all-cause mortality were observed across all three medications: HR = 0.26 (95% CI, 0.24-0.28; p < 0.001) for A-P, HR = 0.33 (0.27-0.40; p < 0.001) for mefloquine, and HR = 0.50 (0.48-0.52; p < 0.001) for N-R. Significant reductions were found for major chronic diseases. Dementia incidence was reduced (HR = 0.48-0.64 across medications), as were diabetes mellitus diagnoses (HR = 0.56-0.75) and diabetes defined by first occurrence of HbA1c ≥ 6.5% (HR = 0.57-0.68). Kidney disease, defined as an eGFR < 45 mL/min/1.73 m², was also significantly reduced (HR = 0.53-0.71). Additional reductions were observed for heart failure (HR = 0.55-0.75), myocardial infarction (HR = 0.55-0.89), hepatic failure (HR = 0.28-0.48), COPD (HR = 0.45-0.69), lung cancer (HR = 0.47-0.69), gastrointestinal malignancies (HR = 0.56-0.70), and brain malignancies (HR = 0.63 for A-P and N-R). All results reached statistical significance (p < 0.05, most < 0.001).

As in the LHS cohort, antiprotozoal exposure was associated with higher risks of specific adverse outcomes, including hearing loss (HR = 1.53-1.65), Sjögren’s syndrome/dry eye (HR = 1.50-2.11), retinal detachment (HR = 1.34-2.01 for A-P and N-R), and lichen planus (HR = 1.59-1.73 for A-P and N-R).

In summary, these consistent patterns across independent systems warranted mechanistic and epidemiologic interpretation, addressed in the Discussion.

## Discussion

In this study, we leveraged comprehensive electronic health records from large-scale healthcare networks in two countries to systematically screen for medications associated with increased human lifespan. Using a medication-wide association approach in a national Israeli cohort, we identified two antiprotozoal agents, A-P and mefloquine, as strongly and significantly associated with reduced all-cause mortality and a broad spectrum of age-related morbidities. These findings were independently validated in the U.S.-based TriNetX network, where consistent reductions in mortality and major age-related outcomes were observed for these medications, as well as for N-R—a medication widely used during the COVID-19 pandemic that contains ritonavir, which has known antiprotozoal activity. Analyses were designed to ensure alignment of time zero between exposed and unexposed groups, anchoring it to exposure status in the exposed cohort and applying established pharmacoepidemiologic principles to avoid immortal-time and other time-related biases.^9,10^

The novelty of this work lies in its approach: by systematically comparing medication histories over a decade in individuals with above-versus below-average longevity within a large population, we identified a class of medications—antiprotozoals—that consistently exhibit associations with increased lifespan and a reduced incidence of common life-shortening conditions, including diabetes and its complications, cardiovascular and cerebrovascular diseases, chronic kidney disease, hepatic failure, chronic obstructive pulmonary disease, dementia, and various malignancies. Replicating these associations across two independent healthcare systems in different countries, and for three mechanistically related but distinct medications, provides convergent evidence for the robustness of these findings.

These results raise the possibility that many age-related conditions with incompletely understood etiologies may, at least in part, be driven by unrecognized, chronic protozoal colonization, which may represent a ubiquitous and overlooked contributor to chronic inflammation, immune dysregulation, and tissue damage underlying the biology of aging. Reduction of protozoal load through short-course antiprotozoal therapy may confer long-term health benefits, as observed in both cohorts.

Remarkably, we also observed consistent increases in the incidence of certain adverse outcomes, including hearing loss, Sjögren’s syndrome^11^, and lichen planus^12^, following antiprotozoal exposure. Rather than undermining the protective associations, these findings provide key mechanistic insights and further strengthen the plausibility of our results. These conditions affect cell types and tissues that are highly sensitive to localized immune-mediated damage, which may arise following the pharmacological elimination of intracellular pathogens. The abrupt clearance of these pathogens could trigger localized inflammation and immune activation at sites of prior intracellular colonization, revealing the pathogen reservoirs while concurrently reducing systemic inflammation.

A key question is which pathogen’s elimination could underlie these associations. Based on global seroprevalence, tissue tropism, and epidemiological links to neurodegenerative and cardiovascular diseases, we hypothesize that *Toxoplasma gondii* is a likely culprit. As an intracellular parasite, *T. gondii* can colonize the gut, disseminate hematogenously, establish low-grade infections in various tissues, including in the CNS, often evading immune detection. Its presence has been associated with increased risks of dementia^13^, schizophrenia^14^, and cardiovascular diseases^15^, while our findings of reduced diabetes and its complications following antiprotozoal treatment align with the hypothesis that eradication of chronic *T. gondii* infection may mitigate systemic inflammation and thromboembolic risks.

*T. gondii* is ubiquitous in human environments, with global seroprevalence estimates ranging from 20% to 80% depending on geography and age. The longer individuals live, the greater their cumulative likelihood of environmental exposure and eventual parasitic colonization, even in the absence of overt clinical disease. As an intracellular parasite, *T. gondii* initially colonizes the gastrointestinal tract, where it may induce chronic low-grade inflammation and promote local immune evasion. By interfering with host cell apoptosis and modulating immune surveillance, *T. gondii* may increase the risk of malignancy. Supporting this, we have shown in a recent study that *T. gondii* DNA signatures are enriched in the stool of individuals with colorectal cancer, indicating active or latent gut colonization in these patients ^16^.

Additionally, *T. gondii* has been reported to colonize the skin^17^, which is anatomically contiguous with the gastrointestinal outlet. While evidence is limited, localized colonization of the skin or mucous membranes occurring more broadly, particularly in aging individuals with declining immune surveillance may contribute to chronic, non-specific inflammation and insulin resistance, a hallmark of diabetes mellitus—a condition we found to be reduced following antiprotozoal treatment. Notably, *T. gondii* infections that remain localized without systemic dissemination may not trigger detectable serological responses, consistent with the parasite’s intracellular lifestyle and immune evasion strategies. Supporting this hypothesis, several studies have reported increased *T. gondii* seroprevalence among individuals with diabetes mellitus ^18,19^.

Furthermore, *T. gondii* is capable of systemic dissemination via hematogenous spread, allowing it to reach and infect a wide range of cell types, notably neuronal tissue within the central nervous system (CNS). Increased seropositivity for *T. gondii* has been associated with a range of neuropsychiatric and neurodegenerative conditions, including dementia and schizophrenia, aligning with our observation of reduced dementia incidence following antiprotozoal treatment ^20–25^.

Parasite dissemination into the systemic circulation may precipitate thromboembolic phenomena, a proposed mechanism for the association of *T. gondii* with cardiovascular and cerebrovascular morbidity ^15,26^. The marked reductions in myocardial infarction and stroke observed following antiprotozoal treatment in our study are consistent with this hypothesis, suggesting that protozoal eradication may mitigate thromboembolic risk by reducing systemic parasite burden and associated vascular inflammation.

Supporting evidence that A-P, mefloquine, and N-R (Paxlovid) possess anti-*Toxoplasma* activity further strengthens the plausibility of our hypothesis.

Atovaquone is a well-established anti-*Toxoplasma* agent that inhibits the parasite’s mitochondrial electron transport chain, impairing its energy production and replication. It is used in combination therapies for toxoplasmosis treatment and prophylaxis, with demonstrated efficacy in suppressing and clearing *T. gondii* infections, including in immunocompromised patients. Proguanil, while primarily targeting *Plasmodium*, may enhance atovaquone’s efficacy by additional mitochondrial inhibition and by inhibiting dihydrofolate reductase (DHFR), thereby interfering with folate metabolism, which is also required for *T. gondii* survival and replication.^27,28^

Mefloquine is an antiprotozoal widely used for malaria prophylaxis and treatment, particularly against *Plasmodium falciparum* strains resistant to chloroquine and other first-line antimalarials. Importantly, mefloquine has demonstrated both in vitro and in vivo activity against *T. gondii*^29,30^. Animal studies have shown reduced parasite loads and extended survival in infected mice treated with mefloquine, indicating its ability to target *T. gondii* tissue cysts and tachyzoites through interference with parasite autophagy and disruption of membrane stability. Additionally, mefloquine is thought to impair parasite autophagic pathways and mitochondrial function, further contributing to its efficacy against intracellular protozoa, including *T. gondii*.

Lastly, N-R (Paxlovid) has not been formally studied for *T. gondii* activity; however, ritonavir and related HIV protease inhibitors have demonstrated *in vitro* anti-*Toxoplasma* activity, likely by interfering with the parasite’s protease systems and replication cycles^31,32^. Furthermore, protease inhibitors have been investigated for synergistic effects with other anti-*Toxoplasma* agents, supporting their potential role in suppressing *T. gondii* replication.

Medications used for malarial prophylaxis are typically administered for the duration of travel (generally several weeks). Combined with their high intracellular penetration, this treatment period may be sufficient to eliminate or clear a substantial portion of the *T. gondii* burden in the host. By reducing parasite load below a critical threshold, such treatments could enable the immune system to prevent the re-expansion of subclinical foci. This mechanism may underlie the durable reductions in chronic inflammation and the lowered risk of age-related diseases that we observed in both populations, persisting for up to a decade after exposure.

In contrast, N-R is administered for only five days to prevent COVID-19 complications—a much shorter course than typical antimalarial regimens—which may limit its effectiveness in eradicating *T. gondii*. Nevertheless, despite this shorter exposure, we observed similar reductions in all-cause mortality and age-related morbidities, along with increased risks of the same auditory, visual, and dermatologic conditions seen with A-P and mefloquine.

The observed associations of antiprotozoal treatment with hearing loss, retinal detachment, Sjögren’s syndrome, and lichen planus further support the biological plausibility of *T. gondii* involvement. These outcomes align with the parasite’s known tissue tropism: congenital toxoplasmosis is a leading cause of sensorineural hearing loss in infants ^33^, and seropositivity has been linked to hearing impairment ^34^; toxoplasma retinitis is the most common cause of posterior uveitis worldwide ^35^; and cutaneous toxoplasmosis frequently occurs in immunocompromised individuals ^17^. Taken together, these findings suggest that the observed increases in localized conditions after antiprotozoal therapy may reflect parasite clearance from its canonical reservoirs, triggering tissue-specific pathology while simultaneously lowering the risk of systemic spread.

Until now, the prevailing view has been that *Toxoplasma gondii* is largely innocuous in immunocompetent humans, aside from its teratogenic effects in newly infected pregnant women and its impact on immunocompromised individuals. Consequently, serological testing is rarely performed outside these contexts. In histopathological examinations, the small, easily overlooked cysts characteristic of *T. gondii* are typically disregarded or unreported due to the absence of a recognized clinical correlation. This likely contributes to the underrecognition of its potential long-term harms and to the absence of therapeutic strategies aimed at parasite clearance in the general population.

### Evolutionary considerations relevant to persistent T. gondii infection

To place these epidemiologic observations in a broader biological context, we outline below a testable evolutionary framework that may help explain the observed patterns.

#### Host death is required for pathogen amplification

Most pathogens gain little by harming their hosts: their success depends on replication and transmission between living hosts, and excessive virulence is usually selected against^36^. *T. gondii* belongs to a distinct ecological class. Its sexual reproduction occurs exclusively in the intestines of felids that consume infected host tissues ^37,38^, a step essential for genetic recombination and for producing resistant oocysts in the massive numbers that enable large-scale environmental spread^39^. Predation or scavenging, both closely coupled to host death, are obligate steps in the parasite’s life cycle and therefore impose strong selection for traits that enable felid consumption of infected tissue ^40^. Loss of aversion to cat odor provides one mechanism by which this process occurs in rodents ^41,42^. However, *T. gondii*’s capacity to spread through a wide variety of host species, including birds, livestock, and humans, is not explained by rodent-specific behavioral interactions. Instead, any physiological, neurological, or systemic impairment that increases vulnerability to predation or scavenging can serve the same reproductive function across host species.

At the same time, felids target prey that provides sufficient biomass, and host impairment occurring before the host has reached optimal edible size or before completion of a critical reproductive window may reduce predation success and endanger reservoir stability. Natural selection should therefore favor parasite traits that increase the host’s susceptibility to feline predation while preserving attainment of adult size and enabling fulfillment of the reproductive phase required for species continuity.

Moreover, host impairments that emerge after reproduction fall into a period when natural selection on the host is weak ^43,44^. Defenses against late-onset pathology confer little reproductive advantage and thus would spread poorly. By concentrating pathogenic effects in this post-reproductive “selection shadow,” a parasite may maximize transmission while minimizing host counter-adaptation.

#### Strategic latency through bradyzoite persistence

After acute infection, *T. gondii* transitions from rapidly replicating tachyzoites to encysted bradyzoites. These cysts persist for decades in long-lived tissues such as neurons and skeletal muscle under immune surveillance^45^. Antigen expression is reduced, and the infection is termed “latent,” a state commonly regarded as innocuous and self-limiting in immunocompetent humans.^38^

However, a metabolically restrained form that selectively encysts in long-lived tissues, actively evades immune elimination, and engages host-specific regulatory and signaling pathways is evolutionarily costly and incompatible with a transmission cycle that ceases with host death. Such elaborate persistence and host-adapted regulation would be unlikely to have evolved across such a wide range of vertebrate hosts unless it contributed to onward transmission. Instead, latency is best understood as an adaptive strategy that allows the parasite to persist in host tissues until the host has aged and fulfilled its reproductive role. At that stage, predation becomes advantageous for parasite transmission, and host debilitation can increase the probability that infected tissues reach a felid without compromising population continuity.

From an evolutionary perspective, latency is therefore unlikely to represent a benign stalemate, but rather a finely adapted transmission strategy that defers pathogenic effects to a stage of host life when such damage can occur without threatening the parasite’s reservoir.

#### Protozoal suppression, mortality and late-life morbidity

In this cohort study, we have identified that exposure to atovaquone–proguanil (A-P) or mefloquine, two widely used malarial prophylactic agents, was associated with large and sustained reductions in all-cause mortality over the subsequent decade in Israel, and replicated the findings in the TriNetX U.S. federated network of approximately 120 million patients, with cohorts rigorously matched on age, sex, calendar time, and major clinical and demographic variables. Consistency across health systems and populations makes simple travel- or demography-based confounding unlikely, particularly in light of the observed balance of major factors of human morbidity.

One parsimonious interpretation of the observed reductions in mortality and morbidity is that these outcomes share a previously underrecognized common substrate: a silent protozoal infection whose virulence is delayed and whose burden was reduced by these antiprotozoal treatments. The observed epidemiologic signals may therefore reflect attenuation of the parasite’s contribution to late-life impairment, a pattern consistent with the evolutionary constraints of *T. gondii*, whose reproductive success is contingent on late-life host impairments that may enable predation or scavenging by felids.

#### Schizophrenia

Schizophrenia is an incompletely understood disease, associated with disordered thought processes and cognitive impairment, for which higher rates of *T. gondii* seropositivity have been detected across multiple populations ^14,20,46,47^, along with immune and metabolic abnormalities^48–52^. If persistent toxoplasmosis contributes to disease risk, two predictions follow: suppression of the parasite should be protective, and impairment of host mechanisms that normally contain intracellular pathogens should increase risk. Both patterns were observed in an Israeli cohort study^53^.

In a medication-wide screen covering all drugs dispensed in the decade before first schizophrenia diagnosis, the strongest protective associations clustered around agents with known anti-*Toxoplasma* activity, including atovaquone–proguanil used for malaria prophylaxis, clindamycin-based regimens used for acne, and moxifloxacin and oxytetracycline/polymyxin-B ophthalmic drops used to treat eye infections or for perioperative prophylaxis. Each of these associations was independently replicated in the TriNetX network with similar effect sizes and directionality. An independent study also reported a protective association for doxycycline, another tetracycline with anti-*Toxoplasma* effects^54^.

Host biological context further supports this interpretation. Before diagnosis, individuals who later developed schizophrenia showed lower vitamin D and T3 levels, higher rates of liver disease and alcoholism, reduced mucosal and barrier integrity, and increased prevalence of chronic viral hepatitis, whereas hepatitis A vaccination and episodes of transient mucosal inflammation were protective. Together, these features indicate impaired immune containment of intracellular parasites ^50^, a milieu in which latent *T. gondii* may shift from quiescence to clinically relevant neurobiological effects.

#### Dementia, viral triggers, and delayed protozoal virulence

*T. gondii* seropositivity is consistently higher across the dementia spectrum, from mild cognitive impairment to Alzheimer’s disease^23,24,55^. If latent toxoplasmosis contributes to late-life neurodegeneration, then interventions that suppress the parasite or prevent its reactivation should reduce dementia risk, whereas conditions that destabilize immune control should increase it.

This pattern has been observed. In an unsupervised medication-wide screen performed in the U.K., a significantly lower incidence of Alzheimer’s disease was detected among individuals prescribed atovaquone–proguanil (A-P) or mefloquine, both CNS-penetrant antiprotozoals ^56^. The protective association with A-P was independently detected in Israel and further replicated in large, age-stratified, rigorously matched U.S. TriNetX cohorts, where protection persisted for more than 10 years after exposure across multiple age strata ^57^.

Independent evidence points to a viral contributing effect. Varicella–zoster virus (VZV) vaccination is associated with reduced dementia risk^58,59^, and herpesvirus genomic signatures are enriched in Alzheimer’s disease brain tissue, while antiviral therapy is associated with lower dementia incidence ^60,61^. Experimental and clinical studies show that herpesvirus infection can impair immune control of latent *T. gondii* and induce protozoal reactivation within the central nervous system^62^.

Consistent with this interaction model, the protective association of A-P was observed mostly for individuals who had not received VZV vaccination, and the protective association of VZV vaccination was observed mostly among those not exposed to A-P, suggesting that both act on the same downstream pathogenic pathway (Figure 2). Longitudinal serologic data further support this framework: individuals seropositive for *T. gondii* had higher subsequent risk of dementia, with serology preceding diagnosis by many years.

Together, medication, vaccination, and serologic evidence converge on a coherent model in which latent protozoal persistence, whose reactivation is enabled by viral coinfection, is associated with heterogeneous late-life neurodegeneration.

#### Cancer

Increased *T. gondii* seropositivity has been observed in both colorectal cancer and glioma in large prospective cohorts, with timing indicating that infection occurs before tumor development^63–65^. If latent toxoplasmosis contributes to carcinogenesis, then protozoal suppression should reduce cancer risk, and this pattern is observed.

In Sweden, use of atovaquone–proguanil (A-P) was associated with reduced colorectal cancer incidence ^66^. This protective association was independently replicated in the TriNetX network, where reduced risk of gastrointestinal malignancies persisted for up to 10 years after exposure and extended to pancreatic cancer^16^. The durability of protection following brief prophylaxis is more consistent with suppression of a persistent carcinogenic factor than with a direct tissue effect occurring only during drug exposure.

Microbiome sequencing experiments provide complementary clues. Reanalysis of a large colorectal cancer metagenomic dataset (PRJEB6070) using protozoa-inclusive reference libraries identified *T. gondii* as the single most discriminatory taxon, exceeding *Fusobacterium*, which was the most discriminatory taxon in the original study ^16^. *T. gondii* DNA was detected in 22.1% of cancer samples compared with 1.53% of controls, with abundance increasing progressively from normal mucosa to adenoma to carcinoma (Figure 3).

Together, prospective serology, durable pharmacoepidemiologic protection, and direct detection of parasite DNA along the adenoma-carcinoma sequence converge on a coherent model in which persistent toxoplasmosis is implicated in malignant transformation in the human gut and possibly beyond.

#### Synthesis and conclusion: a parasite optimized for delayed morbidity

Across independent disease domains, convergent evidence summarized eveals a coherent and internally consistent pattern that suggests *T. gondii* involvement in a wide range of pathogenetic processes.

No currently established host-centered degenerative model readily explains this cross-domain convergence. In contrast, parasite-mediated health deterioration that emerges in late life follows naturally from the evolutionary constraints dictated by *T. gondii*’s life cycle. Because the parasite can complete sexual reproduction only when infected host tissue is consumed by a felid, parasite fitness is inseparable from host death. This ecological constraint would therefore select parasite traits optimized to produce health or behavioral impairment in late adult life, when predation or scavenging becomes advantageous for parasite transmission without endangering population continuity.

In humans, who are frequently and repeatedly exposed to *T. gondii* through environmental oocysts in soil, water, and fresh produce, as well as through consumption of infected meat, seafood, and unpasteurized milk, such parasite-mediated impairment would be expected to manifest as disorders that cluster in later life, including vascular, neurodegenerative, and malignant disease. Long-term tissue persistence, delayed virulence, and the fact that serologic responses may decline or fail to capture low-grade, tissue-restricted infection make it possible that a major contributor to human morbidity could have remained largely unrecognized.

Together, evolutionary logic and human data are consistent with a testable hypothesis: a measurable fraction of late-life morbidity may reflect delayed consequences of *T. gondii* infection rather than purely intrinsic processes. If correct, at least some age-related diseases may depend on targetable infectious factors^67^, with substantial health implications. Based on the epidemiologic observations of this study, we hypothesize that chronic *T. gondii* colonization may contribute to aging-related conditions, thereby explaining the association of antiprotozoal therapies with reduced mortality and morbidity. Our findings raise the possibility that even short courses of antiprotozoal treatment may confer long-term benefits by lowering the burden of T. gondii. These results underscore the need for further research to determine the prevalence of *T. gondii* infection in aging-related diseases and to assess whether targeted clearance strategies—ideally designed to minimize collateral tissue damage—could help mitigate age-associated morbidity and mortality.

## Limitations

Despite the strengths of our large-scale, systematically matched analyses and replication across independent populations, this study has several limitations. First, as an observational study, residual confounding cannot be rigorously excluded. However, the consistent observation of both protective and adverse associations (e.g., hearing loss, Sjögren’s disease, lichen planus) across large cohorts from different countries, for two distinct antimalarials, and in TriNetX also for N-R—a medication administered during COVID-19 primarily to older and less healthy patients and provided at no cost across socioeconomic groups—argues strongly against selection or indication bias as the main explanation.

Second, in the LHS exploratory analysis (Analysis A), individuals exposed to antimalarials did exhibit slightly healthier baseline characteristics, which may explain part, though not all, of the observed risk reduction. In this design, survival status was used to define cases and controls, reflecting its role as an exploratory, hypothesis-generating screen rather than a causal inference framework. While it could in principle amplify baseline differences between groups, these concerns were addressed in Analyses B and C, where group assignment was based directly on first medication exposure (defining the index date) and outcomes were analyzed longitudinally using survival models. In particular, Analysis C further minimized bias through propensity-score matching, which ensured rigorous baseline balance across demographic and clinical factors, including race/ethnicity, BMI, diabetes status, HbA1c, and smoking status.

Finally, while the magnitude of the protective associations is striking, their replication across two independent healthcare systems supports the interpretation that these signals are unlikely to be artifacts of a single study design. The persistence of associations with age-related diseases of distinct etiology and disease course, and across outcomes that often occur in otherwise healthy individuals (such as malignancy and myocardial infarction), further reduces the likelihood of confounding as the primary explanation. Moreover, having constructed the cohorts appropriately and empirically validated the absence of bias through baseline comparisons, observing concordant protective effects together with specific adverse associations (hearing loss, Sjögren’s disease, lichen planus) across drugs with unrelated primary indications provides additional reassurance that the findings are genuine and not primarily attributable to indication bias.

## Conclusion

Overall, the observed protective effects of antiprotozoal medications against mortality and a broad range of aging-associated diseases—together with consistent increases in conditions linked to canonical reservoirs of *Toxoplasma gondii*, observed across two countries and three drugs with distinct indications—generate the hypothesis that chronic protozoal colonization, possibly by *T. gondii*, may be an underrecognized contributor to age-related morbidity. The spectrum of affected outcomes, spanning metabolic, cardiovascular, cerebrovascular, neurodegenerative, and malignant diseases, highlights the potential public health significance of this finding. These results raise the possibility that targeted eradication or suppression of protozoal infections could represent a novel strategy to reduce age-related disease burden and lower mortality. This hypothesis can be evaluated in future studies, including serologic or microbiome-based approaches. Prospective interventional studies may help confirm causality and evaluate therapeutic potential.

## Methods

### Study design

This research was performed in three sequential stages, as illustrated in Figure 1. Each stage was designed to build upon the previous one:

- **Analysis A (Screening stage):** A systematic, medication-wide case-control analysis in a national Israeli cohort to identify candidate medications associated with longevity.
- **Analysis B (Validation-Exploration stage):** A matched cohort study within LHS focused on the candidate antiprotozoal medications identified in Analysis A, aimed at validating associations with mortality and exploring morbidity outcomes.
- **Analysis C (Replication stage):** Independent validation in the U.S.-based TriNetX research network using a matched cohort design, including both the candidate antiprotozoals and nirmatrelvir-ritonavir (N-R), selected as an additional comparator with predicted antiprotozoal properties.

Design and reporting adhere to STROBE and RECORD-PE guidelines for pharmacoepidemiologic cohort studies. At each stage, established pharmacoepidemiologic principles were applied to minimize potential sources of bias, including alignment of time zero and prevention of immortal-time and other time-related biases.^9,10^

### Analysis A: LHS Cohort to Identify Drugs Associated with Increased Longevity

The first stage (Analysis A) was a matched case-control screen designed to identify medications associated with longevity in Leumit Health Services (LHS), one of four nationwide health providers in Israel, delivering comprehensive healthcare to approximately 730,000 members. All Israeli citizens are entitled to comprehensive health insurance and receive a standardized package of healthcare services and medications, as defined by the national “Health Basket” committee. LHS operates a centralized electronic health record (EHR) system, maintaining over two decades of meticulously curated data, including patient demographics, medical diagnoses, healthcare encounters, laboratory test results, and records of prescribed medications. In addition, LHS maintains a comprehensive network of affiliated pharmacies, enabling reliable tracking of medication purchases by its members—including over-the-counter medications—which allows for the identification of associations between medication uptake and clinical outcomes.

Cohort eligibility included current and former LHS members with electronic records of medical visits occurring after age 60. Electronic health records have been available at LHS since 2002; Individuals were eligible only if they had at least 10 years of observable electronic history since their first EHR record. This requirement ensured both cases and controls had comparable longitudinal data availability before cohort entry. Data were extracted from the LHS central data warehouse, encompassing diagnoses and medication purchases through December 31, 2024. Structured query language (SQL) and Python scripts were used to automate data retrieval. Socioeconomic status (SES) was determined based on geocoded residential addresses and classified on a 1-20 scale using the Points Location Intelligence® database, which correlates strongly with official national SES indicators.

Using sex-specific longevity thresholds derived from median life expectancy in our health system—84 years for females and 77 years for males—we constructed two groups of eligible individuals: a high-longevity group, comprising individuals who exceeded the cutoff and were alive at the time of data extraction, and a below-average longevity group, comprising individuals who died before reaching the respective cutoff age. For each individual who died before the threshold, we matched a control with above-average longevity based on sex, birth year, socioeconomic status (SES) group, year of first EHR record in LHS, and year of first ambulatory visit occurring at least 10 years after his first EHR record. Matching prioritized individuals with the closest birth dates and was performed without replacement, such that each control could be matched to only one case. The index date was defined as the ambulatory visit occurring ten years after the first available EHR record for each individual, ensuring comparable duration of electronic medical history prior to index.

### Analysis A: Systemic Screen Procedure

We compared, for each medication, the proportion of individuals with at least one recorded purchase in the 10 years preceding the index date. Medications were identified according to their Anatomical Therapeutic Chemical (ATC) classification, and qualifying purchases were restricted to this fixed 10-year look-back window. For each ATC class, purchase proportions were compared between the above-average and below-average longevity groups using Fisher’s exact test, with false discovery rate (FDR) correction for multiple testing. Signals were defined according to pre-specified criteria prioritizing large effect sizes (odds ratio < 0.5) together with strong statistical support (FDR<0.5%), consistent with a hypothesis-generating screening objective.

### Analysis B: Validation-Exploration Cohort of LHS Patients Exposed to Antiprotozoals

To validate the observed association with reduced mortality and to explore which disease reductions might underlie this effect, we constructed a cohort of individuals aged 40 to 85 who had received at least one prescription for atovaquone-proguanil (A-P) or mefloquine. Each exposed individual was matched to 20 control patients of the same sex, age, socioeconomic status (SES) group, and year of first EHR record, with priority given to controls with the closest birth dates. The index date was defined as the first recorded purchase of the respective medication for the case group, and this exact calendar date was assigned to each matched control. Controls were selected only from individuals confirmed to be alive and active members of LHS at that date, ensuring identical time zero for both groups and preventing immortal time or calendar bias. Matching was performed without replacement, so that each control could be used only once. Because atovaquone-proguanil and mefloquine exposures were rare (≈1% of age-eligible individuals), undocumented use prior to first EHR record would be uncommon and unlikely to materially influence exposure classification.

We compared all-cause mortality and the incidence of age-related conditions using a predefined curated list of 200 ICD-coded medical outcomes routinely monitored at Leumit Research Institute. Time-to-event analyses were conducted using Cox proportional hazards models to estimate hazard ratios (HRs) with 95% confidence intervals. Proportional hazards assumptions were assessed using Schoenfeld residuals. Additional adjustment (age, smoking status, number of ambulatory visits, pregnancy history) was applied in the Cox models, based on the patient status at their assigned index dates. Follow-up was censored at the date of death or the last recorded medical encounter for individuals who remained alive. For each outcome, individuals with a documented diagnosis prior to the index date were excluded from the corresponding analysis.

### Analysis C: TriNetX Validation Cohort

To validate the mortality reduction observed in LHS and to test whether associations could be reproduced in an independent U.S. population, we conducted analyses using TriNetX, a federated research network comprising electronic health record (EHR) data from 70 healthcare organizations and covering more than 120 million patients. Queries were executed online in July 2025.

Validation cohorts included individuals aged 70–89 years at the time of cohort extraction who had either a documented prescription or a qualifying ambulatory visit, which served as the index event. In TriNetX, current age is computed automatically as the time elapsed between the individual’s date of birth and cohort extraction date, irrespective of whether the individual is alive or deceased. For decedents, age is not derived from the date of death; the system abstracts the date of birth and applies the same extraction timestamp used for the cohort query. Accordingly, current age does not condition on survival status and cannot introduce immortal-time bias.

Eligibility additionally required documentation of BMI within the year preceding the index event, ensuring that metabolic matching was based on measurements with comparable recency in all participants. Cases were defined as individuals with a recorded prescription for the medication of interest (atovaquone–proguanil, mefloquine, or nirmatrelvir–ritonavir); controls were individuals with a qualifying ambulatory visit, both within the same BMI-referenced window.

In TriNetX, the index date is automatically defined as the first point at which all inclusion criteria are simultaneously satisfied. For exposed individuals, this necessarily corresponds to their first recorded prescription, and follow-up contributes only after this event; no person-time prior to exposure contributes to the exposed cohort. We used the TriNetX Compare Outcomes pipeline, in which exposed individuals are matched 1:1 to controls using greedy nearest-neighbor propensity-score matching without replacement, ensuring that each control is used only once. Controls were not required to remain unexposed during follow-up, thereby avoiding bias from conditioning on future exposure. TriNetX simply prevents individuals in the exposed cohort from being simultaneously selected as controls, without restricting subsequent medication use. Propensity score matching (PSM) was applied to balance groups on age, sex, ethnicity, race, smoking status, diabetes diagnosis, BMI, and HbA1c levels at baseline. For nirmatrelvir-ritonavir, availability beginning in 2021 resulted in a modest calendar-time shift, which is intrinsic to its recent introduction. Outcomes were compared using Kaplan-Meier survival curves and Cox proportional hazards models to estimate hazard ratios (HRs) with 95% confidence intervals.

Guided by the observed associations in LHS, we evaluated the following outcomes:

- **All-cause mortality**
- **Diabetes** (ICD-10: E08-E13; or incident HbA1c ≥6.5%)
- **Dementia** (F03, G30)
- **Renal insufficiency** (eGFR <45 mL/min/1.73 m²), chosen to ensure a uniform and clinically meaningful definition across healthcare organizations
- **Hepatic failure** (K72)
- **Congestive heart failure (CHF)** (I50)
- **Myocardial infarction (MI)** (I21)
- **Malignancies**: gastrointestinal (C15-C26), urinary tract (C64-C68), brain (C71), lung (C30-C39)
- **Chronic Obstructive Pulmonary Disease (COPD)** (J43-J44)
- **Hearing loss** (H90-H91)
- **Sjögren’s syndrome** (M35.0)
- **Retinal detachment** (H33)
- **Lichen planus** (L43)

These codes were intentionally restricted to ensure uniform diagnostic specificity across healthcare systems, acknowledging reduced sensitivity but minimizing differential misclassification bias. Follow-up was censored at the date of the last recorded medical encounter, up to 4000 days (∼11 years) after the index date. For each outcome, individuals with a documented diagnosis prior to the index date were excluded from the corresponding analysis.

### Data processing in LHS

Data from LHS were extracted from the centralized EHR system and processed using T-SQL queries and Python 3.11 scripts developed by the Leumit Research Institute. To ensure anonymity, all patient identifiers were removed and replaced with study-specific codes prior to analysis. Medication exposure was determined from pharmacy purchase records, covering both prescription and over-the-counter drugs, up to 10 years prior to the index date. Medications were classified according to the Anatomical Therapeutic Chemical (ATC) system.

### Statistical analyses

For LHS data, Fisher’s exact test was used to compare categorical variables, and two-tailed t-tests were applied for continuous variables. Survival between treatment groups was compared using the log-rank test. All tests were two-tailed, with P-values below 0.05 considered statistically significant. Analyses of LHS data were conducted in R version 4.4.

For TriNetX data, analyses were performed within the TriNetX platform using the built-in “compare outcome” analysis pipeline, which applies Kaplan-Meier estimators and Cox proportional hazards models to propensity score matched cohorts.

## Data Availability Statement

The LHS patient-level data generated and analyzed in this study are not publicly available due to patient privacy regulations. Access may be granted to qualified researchers pending approval by the Leumit Health Services Institutional Review Board. Aggregated data supporting the findings of this study are available from the corresponding author upon reasonable request.

TriNetX data are accessible only to licensed member organizations of the TriNetX network, in compliance with HIPAA and institutional guidelines.

## Ethics Statement

The study was reviewed and approved by the Leumit Health Services Institutional Review Board (IRB). Ethical approval was granted, with a waiver of informed consent (LEU-0020-24). All LHS data were analyzed retrospectively and in a fully de-identified manner.

TriNetX data are de-identified and analyzed under agreements with participating health organizations, compliant with HIPAA and institutional guidelines.

## Funding

This study was funded internally by Leumit Health Services. ER was supported in part by the Intramural Research Program, National Institutes of Health, National Cancer Institute, Center for Cancer Research. The content of this publication does not necessarily reflect the views or policies of the Department of Health and Human Services, nor does mention of trade names, commercial products, or organizations imply endorsement by the US Government.

## Conflict of Interest

The authors have declared that there are no conflicts of interest in relation to the subject of this study.

## Use of Artificial Intelligence Tools

During manuscript preparation, the authors used ChatGPT (GPT-4o, OpenAI) for language refinement and style editing. The content, data interpretation, and conclusions are entirely the responsibility of the authors, and the AI tool did not contribute to the scientific analyses or results.

## Author contributions

Conceptualization: AI, EMe, Ema

Methodology: AI, Eme

Investigation: AI, SI, Eme

Writing - original draft: AI

Writing - review & editing: AI, AW, SI, SA, SV, EMa, Eme, ER

## Materials & Correspondence

Correspondence and requests for materials should be addressed to AI

## Conflict of Interest Statement

The Authors declare that there are no conflicts of interest in relation to the subject of this study.

## Abbreviations

A-P: atovaquone-proguanil
BMI: Body Mass Index BP Blood pressure
CHF: Congestive Heart Failure
CI: Confidence Interval
CNS: Central nervous system
COPD: Chronic Obstructive Pulmonary Disorder
EHR: Electronic health records
FDR: False Discovery Rate
GI: Gastrointestinal
GFR: Glomerular Filtration Rate
HbA1c: Hemoglobin A1c
HCO: Healthcare organizations
HR: Hazard Ratio
ICD: International Classification of Diseases
LHS: Leumit Health Services
MI: Myocardial Infarction
N-R: nirmatrelvir-ritonavir
OR: Odds Ratio
PSM: Propensity score matching
SES: Socioeconomic status
SMD: Standardized Mean Difference

## Notes

### Competing Interest Statement

The authors have declared no competing interest.

### Funding Statement

This research was internally funded by Leumit Health Services (LHS). ER was supported in part by the Intramural Research Program, National Institutes of Health, National Cancer Institute, Center for Cancer Research.

### Author Declarations

The study was reviewed and approved by the Leumit Health Services Institutional Review Board (IRB). Ethical approval was granted, with a waiver of informed consent (LEU-0020-24). All LHS data were analyzed retrospectively and in a fully de-identified manner. TriNetX data are de-identified and analyzed under agreements with participating health organizations, compliant with HIPAA and institutional guidelines.

### Summary of Updates

- Expanded the Methods section to provide additional detail on cohort construction, index-date definition, and strategies used to minimize bias, including calendar-time alignment and matching procedures. - Clarified aspects of the statistical analysis and validation strategy, including replication in the TriNetX network. - Expanded the Discussion to contextualize the observed associations and to outline testable, hypothesis-generating interpretations based on existing biological and epidemiologic evidence. - Minor edits throughout the manuscript to improve clarity and precision.

