## Supplementary Tables for "Antiprotozoal medications associated with increased longevity and reduced morbidity in two national cohorts"

**Antiprotozoal Treatment Is Associated with Significantly Lower Mortality and Reduced Age-Related Morbidity**

### Supplementary Table 1. Comparison of medication purchases in the 10 years preceding the index date in the case (shorter lifespan) and control (longer lifespan) groups of the LHS cohort

| **Medication** | **Case**  **N=14,986** | **Control**  **N=14,986** | **Odds Ratio**  **[95% CI]** | **p-value** | **FDR** |
| --- | --- | --- | --- | --- | --- |
| Mefloquine | 11 (0.07%) | 34 (0.23%) | 0.32 [0.15-0.65] | 0.00082 | 0.0001 |
| Atovaquone + Proguanil | 27 (0.18%) | 62 (0.41%) | 0.43 [0.27-0.69] | 0.00026 | 0.0002 |

Counts represent individuals with at least one recorded purchase of the medication in the 10 years preceding the index date. Odds ratios (OR) with 95% confidence intervals (CI) were estimated using Fisher’s exact test, with p-values corrected for multiple testing using the Benjamini–Hochberg false discovery rate (FDR) procedure. The screen evaluated 1,470 distinct compounds; among the highest-ranking associations, the antiprotozoal ATC class P01B (antimalarials) was the only therapeutic category represented by multiple compounds within the top entries, which motivated its selection for downstream validation.
